# The role of the uncinate fasciculus in the risk-architecture of bipolar disorders: A meta-analysis

**DOI:** 10.1101/2021.07.06.21260113

**Authors:** Ellie Xu, Lynn Nguyen, Rebecca Hu, Caitlin M. Stavish, Ellen Leibenluft, Julia O. Linke

## Abstract

**Background:** Bipolar disorder (BD) is a severe mental disorder, characterized by prominent mood swings and emotion regulation (ER) deficits. The uncinate fasciculus (UF), a white matter tract connecting the amygdala and the ventral prefrontal cortex, has been implicated in ER. Aberrancies in UF microstructure may be an endophenotype associated with increased risk for BD. However, findings in individuals with BD and at familial risk for BD (AR) have yielded inconsistent findings. This meta-analysis takes a region-of-interest approach to consolidate the available evidence and elucidate the role of the UF in the risk-architecture of BD.

**Methods:** Using web-based search engines, we identified diffusion tensor imaging studies focusing on the left and right UF and conducted meta-analyses comparing fractional anisotropy (FA) and radial diffusivity (RD) between BD or AR to healthy volunteers (HV).

**Results:** We included 32 studies (n_BD_=1186, n_HV_=2001; n_AR_=289, n_HV_=314). Compared to HV, individuals with BD showed lower FA in the right (p<.0001) and left UF (p=.010), and higher RD in the right UF (p=.009). We found no significant differences between AR and HV. In the right but not the left UF, AR individuals showed higher FA than BD (p=.043).

**Conclusion:** Comparable UF microstructure between AR and HV and higher FA in the right UF in AR compared to BD suggest that aberrancies in UF microstructure is not an endophenotype for BD. Longitudinal studies are needed to determine when UF abnormalities emerge in the context of BD.

## INTRODUCTION

Bipolar disorder (BD) is a severe mental disorder and a leading cause of global disability^1^. To date, the etiology of BD is not well understood, which complicates both its diagnosis^2^ and treatment^3^. Individuals with BD experience episodes of depression and mania, which differ markedly in their emotional and behavioral characteristics^4^. Thus, emotion regulation deficits have been proposed as a relevant mechanism^5,6^. The uncinate fasciculus (UF), a white matter (WM) tract connecting the ventral prefrontal cortex and the amygdala^7^, is thought to play a crucial role in emotion regulation^8–10^. However, reports regarding aberrancies in UF microstructure in individuals with BD and their first-degree relatives, the latter of which are known to be at an increased risk to develop the disorder themselves (AR)^11^, remain inconclusive (**Supplementary Tables S1-S2**).

Here, we present a meta-analysis that extends the literature in three ways. First, we integrate evidence from published reports that used a region-of-interest (ROI) approach to specifically investigate the microstructure of the UF. Prior meta-analyses either included only whole-brain studies, which might be relatively insensitive to effects in a small tract like the UF^12–14^, or focused on data from the ENIGMA consortium (Favre et al., 2019). Second, aside from fractional anisotropy (FA), a diffusion tensor imaging (DTI) metric that has been positively associated with the directionality and coherence of WM tracts^15^, we also investigate radial diffusivity (RD), a metric negatively associated with the myelination of WM tracts^16^. This addition is important as it has been hypothesized that altered myelin plasticity may play a crucial role in the pathogenesis of BD^17^. Third, we include data on individuals AR to test the hypothesis that aberrant microstructure in the UF constitutes an endophenotype for BD^18^.

Endophenotypes are not directly observable features such as brain structure that are (1) heritable, (2) associated with a disorder (in this case, BD) in the population, (3) present regardless of mood state, (4) co-segregate with BD within families, and (5) occur in unaffected relatives at a higher rate than in the general population^19^. The microstructure of the UF is highly heritable, with familial effects explaining 71-80% of variance in FA of the UF^20,21^, but it is unclear whether the remaining endophenotype criteria apply. Prior meta-analyses have consistently reported FA reductions in the genu of the corpus callosum in individuals with BD^12–14,22^; however, findings in the UF have varied, with reports of lower^12,22^ or no FA alterations in the UF in individuals with BD^13,14^. Additionally, previous studies have reported higher, lower, or no FA aberrations in the left or right UF in individuals with BD or AR (**Supplementary Tables S1-S2**).

Altered myelin plasticity has been proposed as a cellular mechanism that contributes to WM abnormalities in BD^17^. While myelin plasticity has not been measured directly in BD, numerous cross-sectional studies investigate RD, a metric associated with the degree of myelination. Specifically, increased RD has been suggested to correspond with decreased myelin integrity^23^. Prior findings include reports of higher RD in the left^24^ or right UF only^25^, bilaterally increased RD^26^, and null findings^24,27^. Thus, results remain inconclusive on whether the degree of UF myelination plays a role in BD.

The considerable heterogeneity of past reports raises the question: which factors contribute to inter-study differences? A prior meta-analysis showed that characteristics of study participants, such as age and sex, explain some inter-study variance in WM findings. For example, studies with a higher proportion of young women with BD report more pronounced FA reductions in WM tracts^13^. Prior meta-analyses also indicate that studies with higher proportions of participants who use lithium or antipsychotics display fewer WM abnormalities in various tracts^13,22^. However, so far, such factors have not been found to contribute to inter-study differences in findings in the UF in particular^13,22^.

Another variable that might influence findings in the UF is the proportion of study participants with a comorbid anxiety disorder. Approximately 60% of individuals with BD meet criteria for a comorbid anxiety disorder^28^, and one mechanism underlying anxiety might be altered UF microstructure^29^. Thus, the number of participants with a comorbid anxiety disorder might explain inter-study differences.

Lastly, prior work found that distinct DTI processing methods, such as tractography and tract-based spatial statistics (TBSS), could lead to false-positive or false-negative results when comparing groups with varying conditions^30^. In our meta-analysis, we examined the processing pipeline as an explanatory variable of inter-study variance.

The primary goal of the present meta-analysis was to determine whether reduced FA in the left and right UF meets the criteria for a BD endophenotype by determining whether it is present in patients with BD independent of current mood state and whether it occurs in at-risk individuals (AR) at a higher rate than in the general population. Further, we aimed to determine whether available evidence supports altered myelination as a mechanism underlying aberrancies in UF microstructure. Finally, we investigated potential sources of heterogeneity in previous findings (i.e., age, sex, lithium and antipsychotic use, comorbid anxiety, and DTI processing pipeline).

## METHODS AND MATERIALS

### Literature Searches

This meta-analysis was first pre-registered in PROSPERO (<u>CRD4202019956</u>). Next, DTI literature in BD published up to October 29th, 2020 was identified using PubMed, EmBase, and WebofScience with the following search terms: white matter AND bipolar disorder. We included studies that 1) compared individuals with BD or AR (the latter defined in the literature as individuals who do not themselves meet criteria for BD, but have a first-degree relative meeting DSM criteria for BD) to healthy volunteers (HV), 2) used an ROI approach that included the UF, and 3) reported means and standard deviations of FA values separately for the left and right UF.

### Data Extraction

We extracted participants’ mean age and sex ratio from each study, as well as mean scores on the Young Mania Rating Scale (YMRS)^31^ (20 studies, n_BD_=777) and Hamilton Depression Rating Scale (HDRS)^32^ (13 studies, n_BD_=536) for individuals with BD. We chose the YMRS and HDRS because these were the most commonly reported measures of mania and depression, respectively. For studies that used a version of the HDRS other than the HDRS-17, we converted mean scores as described in earlier work^13^. Additionally, we extracted the percentage of individuals taking lithium (22 studies, n_BD_=749) or antipsychotic medication (22 studies, n_BD_=798), the percentage of individuals with a lifetime comorbid anxiety disorder (20 studies, n_BD_=619), and the DTI processing pipeline used (28 studies, n=3187) from each study that provided this information in the published manuscript. We did not exclude any studies that had missing information on mean YMRS and HDRS scores, lithium or antipsychotic medication use, or comorbid anxiety disorders. All studies provided information on mean age, sex ratio, and the DTI processing pipeline. Given our hypotheses, we obtained means and standard deviations of FA values of the left and right UF separately for all study groups. We did not extract mania and depression scores and medication status of individuals AR. Of the 11 studies on individuals AR, only five reported a YMRS mean score, three reported a HDRS mean score, and six reported medication status. Seven of the 11 studies included only asymptomatic individuals AR, while four were conducted in symptomatic individuals AR.

Missing information was requested from the corresponding authors of 33 studies. We obtained missing data from 13 studies^26,27,33–43^. Using the Newcastle-Ottawa assessment scale (NOS), we evaluated the quality of all studies based on how the study groups were selected, how comparable they were, and how their DTI metrics were ascertained^44^ (**Supplementary Table S3**). All data were initially extracted by EX and RH and cross-checked independently by LN.

### Data Analyses

All analyses were conducted using the metafor package (version 2.4-0) for R software (R Foundation for Statistical Computing, Vienna, Austria; http://www.r-project.org/). First, we calculated the effect sizes for each study as standardized mean difference values (Cohen’s d). Next, we used the effect sizes to conduct random-effects inverse-variance weighted meta-analyses.

We tested whether lower FA in the left and right UF is an endophenotype of BD by (1) comparing individuals with BD and HV (2nd criterion), (2) conducting a meta-regression with YMRS- and HDRS-scores as predictors to test independence of mood state (3rd criterion), and (3) comparing individuals AR and HV (5th criterion). In addition, we also conducted a meta-analytic comparison between individuals AR and individuals with BD as the absence or presence of such differences could provide additional evidence, although not proof, for or against the endophenotype hypothesis in the event that AR and HV do not differ. Lastly, we compared RD of the left and right UF between individuals with BD and HV to elucidate whether altered UF myelination plays a role in BD. We did not compare RD between individuals AR and HV given the limited number of reports (n=4). Effects were considered significant at p < 0.05.

We assessed publication bias using Egger’s test for asymmetry. Studies that were identified as outliers using the Egger’s test were removed from the analysis reported in the main body of the manuscript. However, analyses including the outliers are shown in the supplement. Robustness of results were ensured with jackknife sensitivity analyses.

To determine potential effects of demographic and clinical characteristics on heterogeneity in DTI findings, we conducted meta-regression analyses. Specifically, we examined the effect of age, sex, lithium and antipsychotic use, comorbid anxiety, and DTI processing pipeline. These analyses were considered significant at p < 0.008, using Bonferroni correction for multiple comparisons. Given that meta-regression analyses can only be expected to produce robust results when at least 20 studies are included^45^, we examined the effect of these variables on UF FA only for studies comparing BD to HV (n=28), but not studies comparing RD between BD and HV (n=8) or studies comparing FA between AR and HV (n=11). Extracted data and code are provided on https://osf.io/rdcpf/.

## RESULTS

### Literature Search and Quality Assessment

In the initial literature search, we identified 3970 studies (**Figure 1**). We screened the titles and abstracts and selected 160 studies for full-text review. Studies were excluded if they did not include the UF as a ROI, used only whole-brain analyses, were missing FA data, or did not meet the NOS quality assessment criterion (score > 5) (**Figure 1, Supplementary Table S4**). Thirty-two distinct studies met these criteria and were included in this meta-analysis. Overall sample characteristics are shown in **Table 1**, and details of individual studies are provided in **Supplementary Tables S1-S2**.

**Figure 1.**
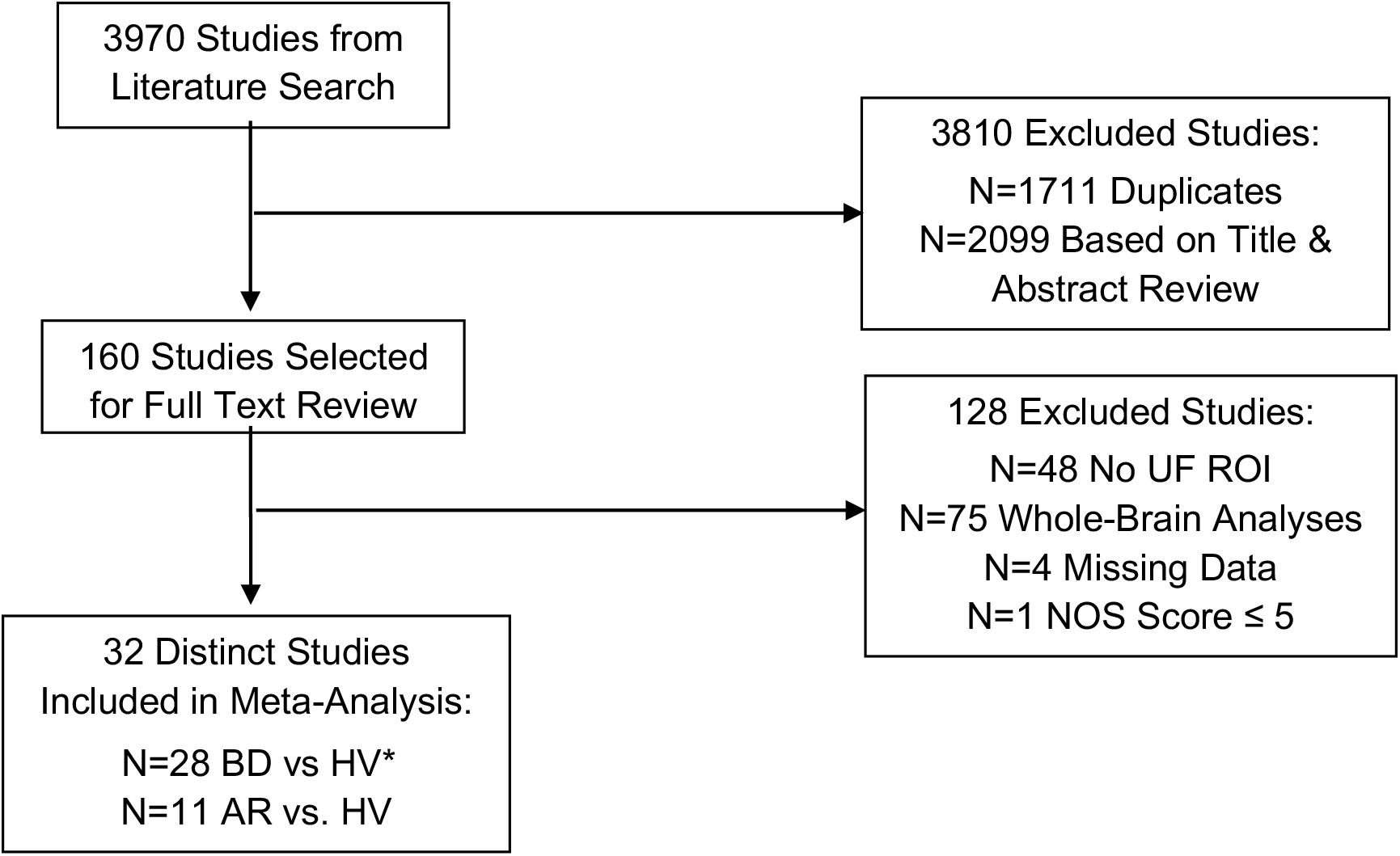
Flow chart of the literature search process. **Abbreviations: AR**, at-risk relatives; **BD**, bipolar disorder; **FA**, fractional anisotropy; **HV**, healthy volunteers; **ROI**, region-of-interest; **UF**, uncinate fasciculus; **NOS**, Newcastle-Ottawa Assessment Scale. **\*** Final study count does not include one outlier.

**Table 1.** Overall sample characteristics for the meta-analyses of fractional anisotropy and radial diffusivity data.

|  | BD vs. HV |  | AR vs. HV |
| --- | --- | --- | --- |
|  | FA | RD | FA |
| **Number of studies** | 28 | 8 | 11 |
| **Total sample size** | 3187 | 1687 | 603 |
| **BD or AR sample size** | 1186 | 411 | 289 |
| **Mean Age (Years)** | 33.5 | 33.9 | 26.1 |
| **Female (%)** | 51.7 | 50.3 | 49.3 |
| **Illness Duration (Years)** | 13.1 | 13.5* | - |
| **Medicated (%)** | 76.6* | 85.6* | 0.1** |
| **Anxiety (%)** | 17.8* | 20.9 | 7.2 |
| **Lithium (%)** | 38 | 36.9 | 0.0** |
| **Antipsychotics (%)** | 40.2 | 41.2 | 0.0** |
| **Mean YMRS** | 3.0* | 4.1* | 0.8** |
| **Mean HDRS-17** | 5.6** | 3.1** | 0.5** |

**Abbreviations:** AR, at-risk relatives; BD, bipolar disorder; FA, fractional anisotropy; HDRS-17, Hamilton Depression Rating Scale, 17-item; HV, healthy volunteers; RD, radial diffusivity; YMRS, Young Mania Rating Scale.
\* These means are based on less than 75% of studies, which reported this information in the published manuscript.
\*\* These means are based on less than 50% of studies, which reported this information in the published manuscript.

### Is Aberrant Fractional Anisotropy in the Uncinate Fasciculus an Endophenotype for Bipolar Disorder?

The Egger’s test indicated one outlier that showed greatly reduced FA of the left and right UF in individuals with BD^46^ (**Supplementary Figures S1-S3)**. After removing this study, the Egger’s test indicated no publication bias for FA in the left (z = 0.77, p = 0.442) or right UF (z = 0.08, p = 0.938). We observed lower FA in the left and right UF in individuals with BD compared to HV (**Figure 2**). The estimated weighted mean difference (WMD) in FA was -0.21 (95% CI = [-0.37, -0.05], p = 0.010; **Figure 3**) in the left and -0.31 (95% CI = [-0.43, -0.19], p < 0.0001; **Figure 4**) in the right UF. Jackknife sensitivity analyses found that no single study drove these results.

**Figure 2.**
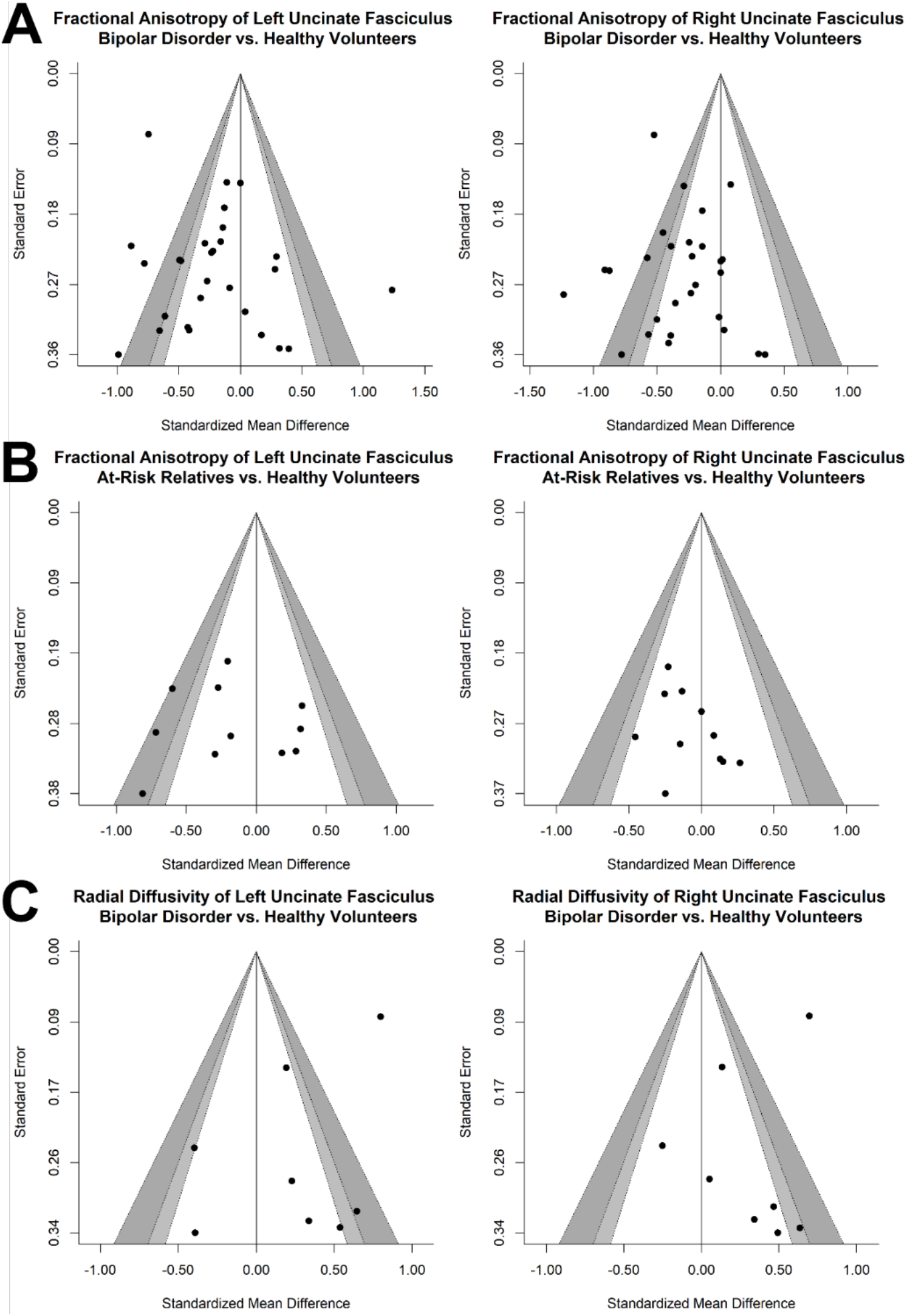
Funnel plots depicting standardized mean differences of fractional anisotropy (FA) and radial diffusivity (RD) between study groups. Funnel plots are scatter plots that represent each study’s reported effect size (x-axis) plotted against its size (y-axis). Asymmetric funnel plots may suggest publication bias. **(A)** Funnel plot of FA in the left and right uncinate fasciculus (UF) of individuals with bipolar disorder (BD) versus healthy volunteers (HV). **(B)** Funnel plot of FA in the left and right UF of at-risk relatives (AR) versus HV. **(C)** Funnel plot of RD in the left and right UF of BD versus HV groups.

**Figure 3.**
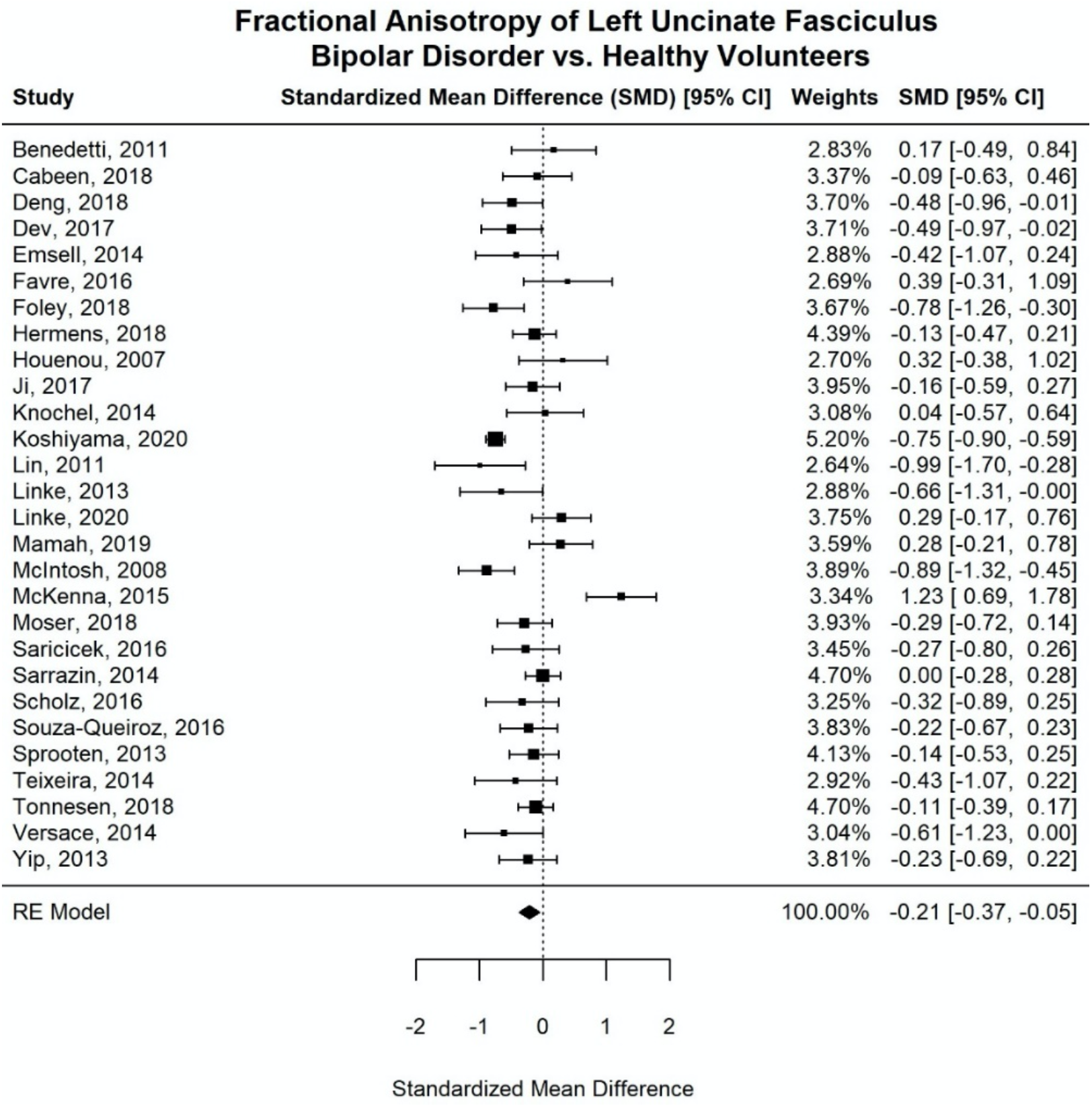
Forest plot of fractional anisotropy in the left uncinate fasciculus of individuals with bipolar disorder versus healthy volunteers. **Abbreviations: CI**, confidence interval; **RE model**, random-effect model.

**Figure 4.**
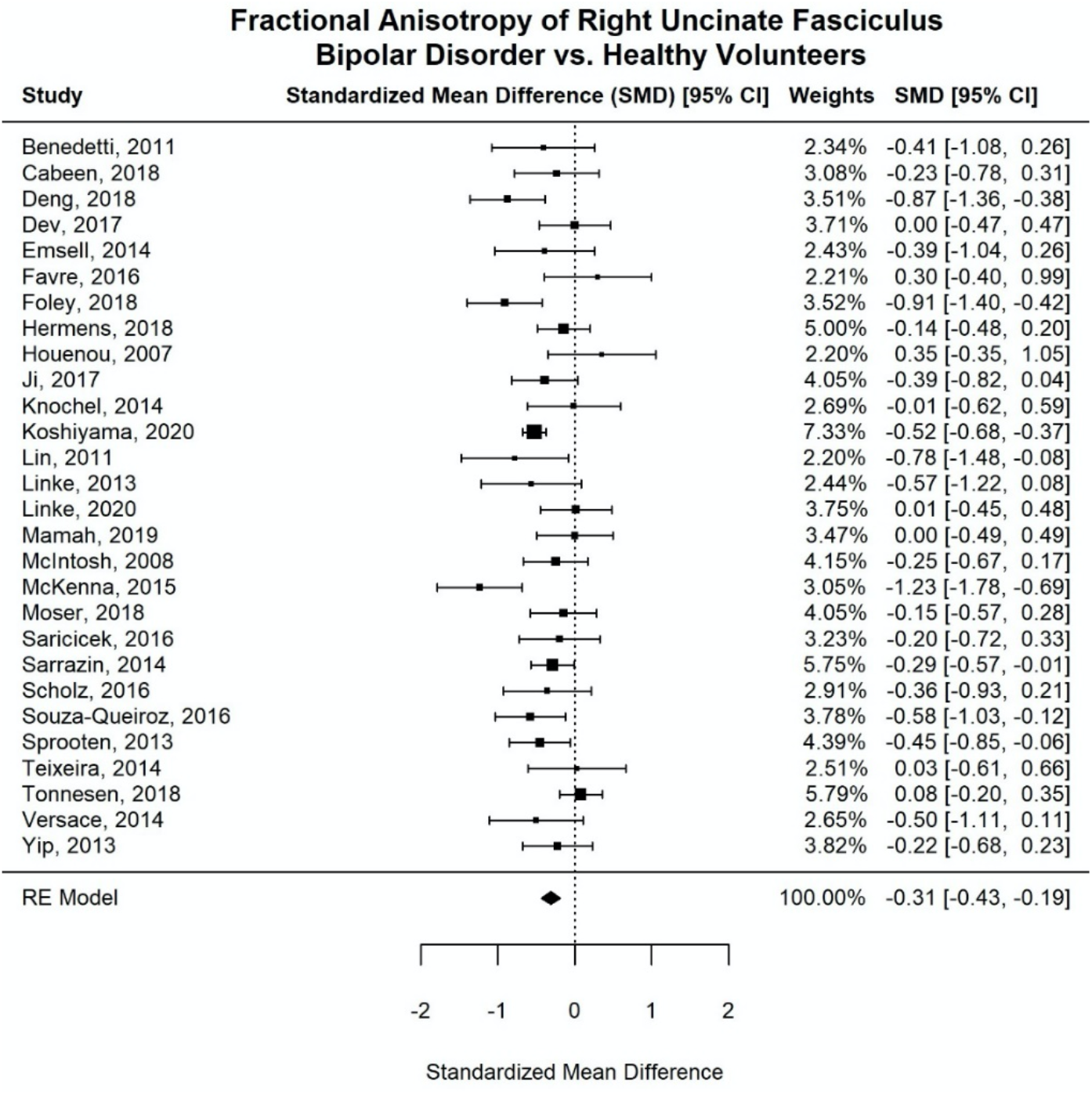
Forest plot of fractional anisotropy in the right uncinate fasciculus of individuals with bipolar disorder versus healthy volunteers. **Abbreviations: CI**, confidence interval; **RE model**, random-effect model.

Elevated mood did not predict FA in the left (p = 0.721) or right UF (p = 0.232). Depressed mood was also not a predictor of FA in the left (p = 0.879) or right UF (p = 0.623). Therefore, altered FA of the UF in individuals with BD appears to be state-independent.

For the comparison between individuals AR to HV, the Egger’s test indicated the absence of a publication bias for FA in the left (z = -0.02, p = 0.981) or right UF (z = 1.00, p = 0.316), leading us to include all 11 studies in the analysis. FA in the UF was comparable between individuals AR and HVs in the left (p = 0.119; **Supplementary Figure S4**) and right UF (p = 0.209; **Supplementary Figure S5**). Further, individuals AR showed higher FA than individuals with BD in the right UF (p = 0.043), but not the left UF (p = 0.786).

### Does Radial Diffusivity in the Uncinate Fasciculus Differ for Individuals with Bipolar Disorder?

The Egger’s test suggested the absence of a publication bias among the studies that reported RD (left UF: z = -1.17, p = 0.244; right UF: z = -0.51, p = 0.613). Thus, all studies were included in the final analysis. Comparing individuals with BD to HV, we observed higher RD in the right UF (WMD = 0.32, 95% CI = [0.08, 0.57], p = 0.009; **Figures 2, 5**). There was no difference in RD in the left UF (p = 0.079; **Supplementary Figure S6**).

**Figure 5.**
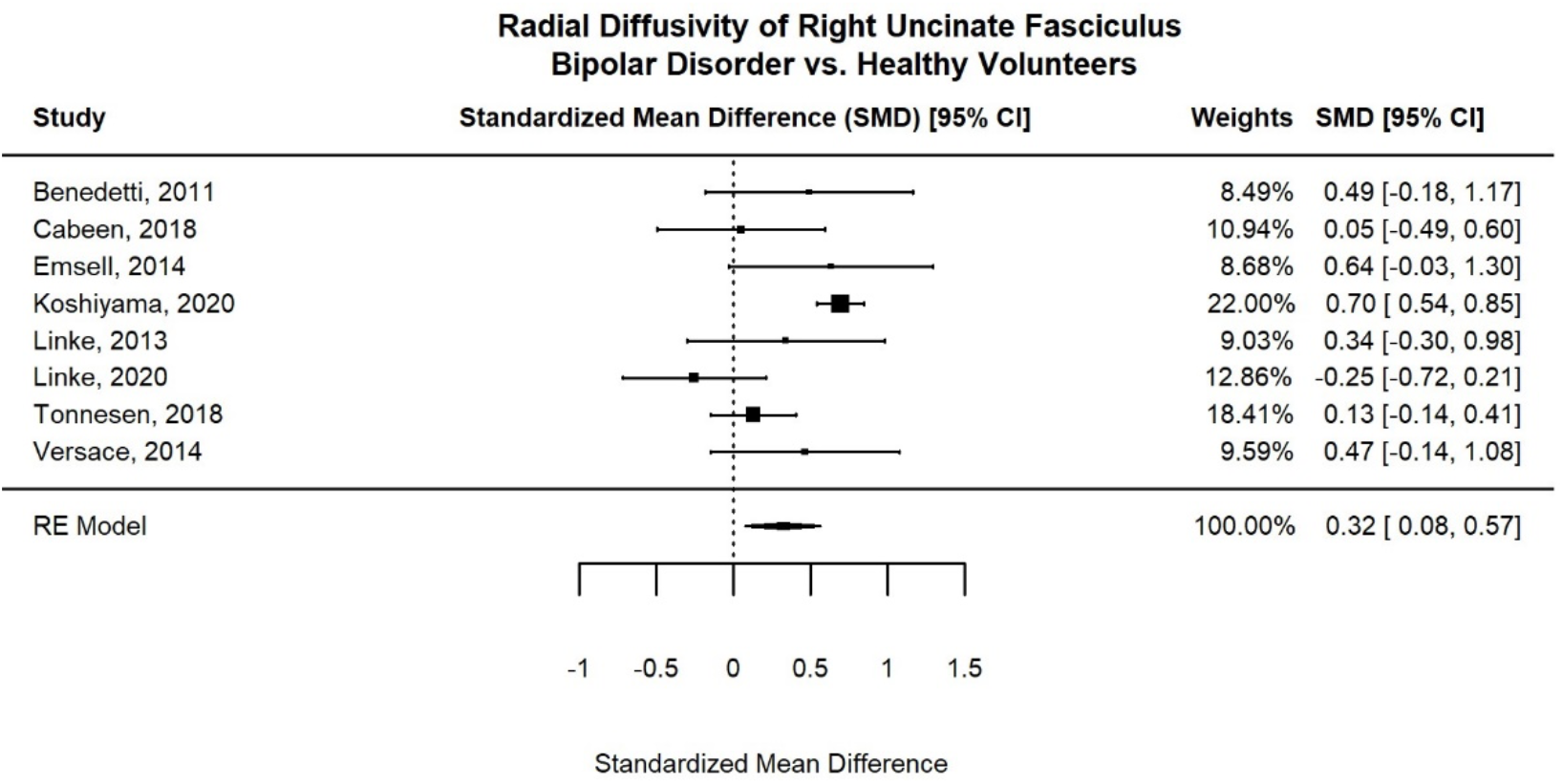
Forest plot of radial diffusivity in the right uncinate fasciculus of individuals with bipolar disorder versus healthy volunteers. **Abbreviations: CI**, confidence interval; **RE model**, random-effect model.

### Do Demographic and Clinical Characteristics Explain Inter-study Heterogeneity?

None of the examined variables (age, sex, lithium and antipsychotic use, comorbid anxiety, or DTI processing pipeline) influenced heterogeneity in UF findings across studies in individuals with BD (all *p*s_*uncorrected*_ > 0.115) (**Supplementary Table S5**).

## DISCUSSION

This meta-analysis associates BD with reduced FA in the left and right UF. This effect was observable independent of mood state at the time of scanning. However, contrary to the hypothesis that abnormal microstructure of the UF is an endophenotype of BD, no consistent differences between individuals AR and HV were observed. In addition, we show evidence that abnormalities in the right UF relate to aberrant myelination as RD values were higher in individuals with BD compared to HV. Finally, differences between studies could not be explained by age, sex ratio, the proportion of participants using lithium or antipsychotics, the proportion of participants with a lifetime comorbid anxiety disorder, or the DTI processing pipeline.

### Is Aberrant Fractional Anisotropy in the Uncinate Fasciculus an Endophenotype for Bipolar Disorder?

Our finding of lower FA in the UF in BD compared to HV is consistent with a previous meta-analysis of ENIGMA data that also employed a ROI approach^22^. The UF has been implicated in emotion regulation; specifically, higher FA in the UF has been suggested to facilitate amygdala regulation and has been positively associated with the use of reappraisal^8–10^. Consistent with previous studies showing perturbed emotion regulation even in euthymic patients^47^, this meta-analysis found altered FA in the UF to be associated with BD regardless of current mood state. This finding suggests that lower FA in the UF is not secondary to an affective episode in individuals with BD.

Notably, we found no FA differences between individuals AR and HV and significant differences in the effect between individuals AR vs. HV and individuals with BD vs. HV in the right UF. These two findings suggest that altered microstructure in the right UF is not an endophenotype for BD. Our findings in the left UF were more inconclusive as our findings indicate that the effect between individuals AR vs. HV and individuals with BD vs. HV is comparable. Prior work has suggested that individuals AR display more subtle WM aberrancies than individuals with BD^40^. It is possible that the present meta-analysis, which included only eleven studies in individuals AR, was insufficiently powered to detect an effect. Prior work has shown that, though FA in the UF does not differ between individuals AR and HV at baseline, lower FA in the right UF predicts BD onset in AR youth over a follow-up period of six years^48^. Thus, it is possible that reduced FA in the right UF may be specific to individuals AR in the prodromal stage of BD. More studies examining WM microstructure in individuals AR, ideally in a longitudinal design that reveals whether abnormalities are prospectively associated with the onset of the disorder, are warranted to replicate earlier findings^48^ in larger samples.

### Does Radial Diffusivity in the Uncinate Fasciculus Differ for Individuals with Bipolar Disorder?

We observed higher RD in individuals with BD compared to HV, supporting the hypothesis that aberrant myelination might be a mechanism in BD^17^. Overall, the literature on myelination and myelin plasticity in BD is nascent. Aberrancies in myelin plasticity may reflect responses to environmental stimuli^49^, and/ or perturbations in developmental myelination. Future studies should explore whether these are relevant mechanisms in BD. Also, what are the cellular mechanisms that underlie alterations in the myelination of WM tracts in BD? Future studies addressing these questions are necessary to better understand the role of myelination in the pathophysiology of BD.

### Do Demographic and Clinical Characteristics Explain Inter-study Heterogeneity?

Consistent with previous meta-analyses^13,22^, we did not find an association between lithium or antipsychotic use and FA in the UF. This provides further evidence that the effect of these two types of psychotropic medication is circuit-specific, and that these circuitry do not entail the UF. In contrast to prior meta-analyses^13,22^, age and sex did not explain heterogeneity in UF findings across studies. Further, differences in the DTI processing pipeline did not explain variability of the findings. Finally, we show that aberrancies in the UF in BD are not driven by comorbid anxiety disorders. However, future studies replicating this finding are warranted, as the subset of studies that entered this analysis was small (20 studies, n_BD_=619).

## Limitations

Some subsamples were small, and these findings should be interpreted cautiously. Specifically, we refer to the comparison between individuals AR and HV (11 studies, n=603), and the comparison of RD between individuals with BD and HV (8 studies, n=1687). Further, of the studies that reported mood state of individuals with BD, most studies comprised euthymic individuals (n=608). Finally, studies including individuals AR were very heterogeneous; four included relatives who were symptomatic, whereas seven did not. There was also variability regarding the familial relationship between the individual AR and the index case (e.g., offspring vs. sibling). Given the small sample size, we were unable to investigate effects of these variables on FA in the UF.

## Conclusion

This meta-analysis confirms lower FA of the UF in individuals with BD, but suggests that these alterations may not represent an endophenotype for BD because they were not observable in individuals AR that also showed higher FA in the right UF compared to BD. However, considering the small number of studies in individuals AR and the fact that results remain inconclusive with regard to the left UF, more studies investigating WM in AR individuals are warranted. Finally, individuals with BD exhibited increased RD in the right UF, providing evidence for the hypotheses that altered myelin plasticity is a mechanism in BD.

## Data Availability

All meta-data and R-code can be accessed via the Open Science Forum (OSF).

https://osf.io/rdcpf/

## ACKNOWLEDGMENTS

This work was supported by the Intramural Research Program of the NIMH (ZIA: MH002778-18).

