## Supplementary Material for "The role of the uncinate fasciculus in the risk-architecture of bipolar disorders: A meta-analysis"

***Supplemental Information***

Prior to removing the one outlier[17], we observed lower fractional anisotropy (FA) in the left and right uncinate fasciculus (UF) in individuals with bipolar disorder compared to healthy volunteers (**Supplementary Figures S1-S3**). The estimated weighted mean difference in FA was -0.27 (95% CI = [-0.47, -0.07], $p = 0.009$; **Supplementary Figure S2**) in the left and -0.36 (95% CI = [-0.51, -0.21], $p < 0.0001$; **Supplementary Figure S3**) in the right UF. Jackknife sensitivity analysis found that no single study drove these results for either the left or right UF. However, Egger's test revealed significant funnel plot asymmetry in the left ($z = -4.33$, $p < 0.0001$) and right UF ($z = -2.76$, $p = 0.006$).

**Table S1.** Characteristics of bipolar disorder studies included in the region-of-interest meta-analyses. All studies reported at least fractional anisotropy data, and a subset also reported radial diffusivity data. HDRS was converted to an estimated 17-item score if the 21- or 25-item scales were used.

| | | | Individuals with Bipolar Disorder | | | | | | | | | | | Healthy Volunteers | | |
|---|---|---|---|---|---|---|---|---|---|---|---|---|---|---|---|---|
| Study | **DTI Processing Pipeline** | **RD** | ***n*** | **Mean Age (Years)** | **Female (%)** | **Illness Duration (Years)** | **Subtype** | **Mood State** | **YMRS** | **HDRS-17** | **Comorbid Anxiety (%)** | **Lithium Use (%)** | **Antipsychotic Use (%)** | ***n*** | **Mean Age (Years)** | **Female (%)** |
| Benedetti 2011 [1] | Tractography | X | 15 | 48.4 | 66.7 | 12.4 | BD-I | Euthymic | - | - | 0.0 | 53.5 | 40.0 | 21 | 46.4 | 47.6 |
| Cabeen 2018 [3] | Tractography | X | 26 | 13.9 | 30.8 | - | BD-I | 69% Euthymic, 11.5% Depressed, 8% Elevated | 7.3 | - | 19.2 | 31.0 | 60.0 | 26 | 13.9 | 34.6 |
| Deng 2018 [4] | Tractography | | 29 | 25.7 | 27.6 | 3.9 | - | Depressed | 2.3 | 20.3 | - | 0.0 | 0.0 | 44 | 28.3 | 38.6 |
| Dev 2017 [5] | TBSS | | 33 | 46.3 | 73.0 | - | - | Euthymic | 1.18 | 4.50 | 0.0 | - | - | 38 | 48.8 | 55.0 |
| Emsell 2014 [6] | Tractography | X | 19 | 43.3 | 52.6 | 15.6 | BD-I | Euthymic | - | - | 5.3 | 47.4 | 15.8 | 18 | 41.7 | 44.4 |
| Favre 2016 [7] | Tractography | | 24 | 45.2 | 50.0 | 17.8 | - | Euthymic | 2.79 | - | 0.0 | 47.0 | 9.0 | 12 | 43.6 | 66.7 |
| Foley 2018 [8] | Tractography | | 32 | 45 | 68.0 | 26.0 | BD-I | Euthymic | 2.72 | 3.50 | 31.0 | 25.0 | 63.0 | 40 | 43.5 | 60.0 |
| Hermens 2018 [9] | TBSS | | 76 | 22.2 | 76.3 | 7.0 | - | 59.2% Euthymic, 22.4% Depressed, 14.5% Elevated | 6.00 | 11.7 | 35.0 | - | 55.3 | 59 | 23.8 | 64.4 |
| Houenou 2007 [10] | Tractography | | 16 | 41.9 | 50.0 | 23.8 | - | Euthymic | 0.69 | 1.60 | 6.3 | 87.5 | 56.3 | 16 | 40.5 | 43.8 |
| Ji 2017 [11] | Tractography | | 72 | 26.2 | 61.1 | 8.6 | BD-I | 91.7% Euthymic, 8.3% Elevated | 4.88 | - | 45.8 | - | - | 30 | 26.1 | 46.7 |
| Knochel 2014 [12] | TBSS | | 21 | 35.7 | 42.9 | 7.6 | BD-I | - | - | - | 0.0 | - | - | 21 | 37.0 | 38.1 |
| Koshiyama 2020 [13] | TBSS | X | 211 | 45.7 | 50.2 | - | - | - | - | - | - | - | - | 844 | 36.7 | 46.0 |
| Lin 2011 [14] | Tractography | | 18 | 28.5 | 66.7 | 1.3 | - | 11.1% Euthymic, 88.9% Depressed | - | - | 0.0 | 61.1 | - | 16 | 29.9 | 75.0 |
| Linke 2013 [15] | TBSS | X | 19 | 45.0 | 57.9 | 15.4 | BD-I | Euthymic | 0.90 | 0.70 | 10.5 | 31.6 | 0.0 | 19 | 45.0 | 57.9 |
| Linke 2020 [16] | Tractography | X | 36 | 16.8 | 47.2 | - | - | 86.1% Euthymic, 12.5% Depressed, 1.4% Elevated | 5.90 | - | 44.4 | 33.3 | 50.0 | 36 | 15.0 | 55.6 |
| Mahapatra 2017 [17] | Tractography | | 16 | 29.8 | 43.8 | 3.8 | BD-I | Euthymic | 4.63 | 6.30 | 0.0 | 37.5 | 75.0 | 15 | 29.8 | 40.0 |
| Mamah 2019 [18] | TBSS | | 33 | 26.5 | 45.5 | 9.9 | BD-I | - | 3.50 | - | - | 18.2 | - | 30 | 24.5 | 50.0 |
| McIntosh 2008 [19] | Tractography | | 40 | 39.9 | 47.5 | 18.8 | - | - | 0.60 | 2.40 | - | 55.0 | 45.0 | 49 | 35.3 | 42.9 |
| McKenna 2015 [20] | TBSS | | 26 | 45.2 | 69.0 | 25.9 | BD-I | Euthymic | 1.23 | 3.20 | 0.0 | 31.0 | 54.0 | 36 | 46.3 | 53.0 |
| Moser 2018 [21] | TBSS | | 37 | 27.2 | 32.4 | 6.3 | BD-I | - | - | - | - | 40.5 | 81.1 | 48 | 29.8 | 41.7 |
| Saricicek 2016 [22] | TBSS | | 27 | 34.8 | 59.3 | 10.3 | BD-I | Euthymic | - | - | 0.0 | 59.3 | 59.3 | 29 | 33.7 | 58.6 |
| Sarrazin 2014 [23] | Tractography | | 118 | 36.3 | 60.2 | 15.6 | BD-I | 67.8% Euthymic, 3.8% Depressed, 28.4% Elevated | 2.58 | 9.90 | - | 33.1 | 44.1 | 86 | 37.3 | 52.3 |
| Scholz 2016 [24] | TBSS | | 24 | 44.0 | 41.7 | 21.9 | BD-I | Euthymic | 0.63 | 1.20 | 0.0 | 50.0 | 0.0 | 24 | 44.0 | 41.7 |
| Souza-Queiroz 2016 [26] | Tractography | | 32 | 35.8 | 37.5 | 15.5 | - | - | 3.90 | - | 15.0 | 89.0 | 93.0 | 47 | 36.4 | 53.2 |
| Sprooten 2013 [27] | TBSS | | 61 | 31.7 | 71.9 | 10.0 | BD-I | Euthymic | 1.00 | 2.00 | 46.9 | 18.8 | 35.9 | 43 | 30.1 | 67.4 |
| Teixeira 2014 [28] | TBSS | | 18 | 12.3 | 33.3 | 2.8 | - | 44.4% Euthymic, 11.1% Depressed, 44.4% Elevated | 8.70 | - | 50.0 | 0.0 | 0.0 | 20 | 12.7 | 33.3 |
| Tonnesen 2018 [29] | TBSS | X | 61 | 31.7 | 52.5 | 10.2 | - | - | - | - | - | 25.0 | 64.6 | 293 | 31.9 | 41.3 |
| Versace 2014 [31] | Tractography | X | 24 | 33.2 | 66.7 | 13.8 | BD-I | Euthymic | 2.30 | 5.60 | 45.8 | - | 58.3 | 19 | 34.0 | 52.6 |
| Yip 2013 [33] | TBSS | | 38 | 20.9 | 47.0 | - | BD-II | Euthymic | 1.80 | 6.10 | - | 0.0 | 0.0 | 37 | 21.2 | 46.0 |

**Abbreviations: BD-I,** bipolar disorder type I; **BD-II,** bipolar disorder type II; **HDRS-17,** Hamilton Depression Rating Scale, 17-item; **RD,** radial diffusivity; **YMRS,** Young Mania Rating Scale.

**Table S2.** Characteristics of studies comparing individuals at-risk for bipolar disorder with healthy volunteers. All studies reported fractional anisotropy data, and a subset also reported radial diffusivity data.

| Study | At-Risk Relatives | | | | | Healthy Volunteers | | |
|---|---|---|---|---|---|---|---|---|
| | ***n*** | **Index Relationship** | **Index BD Type** | **Mean Age (Years)** | **Female (%)** | ***n*** | **Mean Age (Years)** | **Female (%)** |
| Besenek 2019 [2] | 19 | Offspring | BD-I, BD-II | 14.3 | 47.4 | 19 | 17.4 | 31.6 |
| Emsell 2014 [6] | 21 | Not Specified[1] | BD-I | 42.5 | 42.9 | 18 | 41.7 | 44.4 |
| Foley 2018 [8] | 17 | Siblings | BD-I, BD-II | 47.2 | 53.0 | 40 | 43.5 | 60.0 |
| Linke 2013 [15] | 22 | Not Specified[1] | BD-I | 28.0 | 50.0 | 22 | 28.0 | 50.0 |
| Linke 2020 [16] | 36 | Offspring, Siblings | BD-I, BD-II | 14.0 | 47.2 | 36 | 15.0 | 55.6 |
| Mahapatra 2017 [17] | 15 | Not Specified[1] | BD-I | 30.9 | 40.0 | 15 | 29.8 | 40.0 |
| Saricicek 2016 [22] | 20 | Siblings | BD-I | 36.3 | 50.0 | 29 | 33.7 | 58.6 |
| Shakeel 2020 [25] | 27 | Not Specified[1] | BD-NOS | 18.5 | 63.3 | 33 | 19.1 | 50.0 |
| Sprooten 2013 [27] | 59 | Siblings | BD-I | 30.4 | 60.0 | 43 | 30.1 | 67.4 |
| Teixeira 2014 [28] | 18 | Offspring | BD-I | 12.7 | 50.0 | 20 | 12.7 | 33.3 |
| Versace 2018 [32] | 35 | Offspring | BD-NOS | 13.7 | 48.6 | 39 | 14.0 | 41.0 |

**Abbreviations: BD-I**, bipolar disorder type I; **BD-II**, bipolar disorder type II; **BD-NOS**, bipolar disorder not otherwise specified.

**1** At-risk individuals were unaffected individuals with a first-degree relative with bipolar disorder.

**Table S3.** Newcastle-Ottawa Quality Assessment Scale (Adapted for this Meta-Analysis). A study can be awarded a maximum of one point for each numbered item within the Selection and Exposure categories. A maximum of two points can be given for Comparability.

| | | |
|---|---|---|
| **Selection** | 1) Is the case definition (diagnosis of BD) adequate? | a) **1 point: clinician interview was conducted** (e.g., SCID, K-SADs)<br>b) **0 points**: chart review or self-report |
| | 2) Representativeness of cases | a) **1 point: broad inclusion criteria,** e.g., *all* youth or *all* adults with BD, or patients with BD identified from inpatient and outpatient visits<br>b) **0 points**: restrictive exclusion criteria, e.g., only age 60+ adults with BD, only patients with first-time mania, only unmedicated patients with BD, only patients with BD taking lithium, or only patients with BD with no other psychiatric comorbidities |
| | 3) Selection of controls | a) **1 point: community controls**<br>b) **0 points:** hospital controls or no description |
| | 4) Definition of controls | a) **1 point: no history of bipolar or other psychiatric disorders**<br>b) **0 points:** no description |
| **Comparability** | 1) Comparability of cases and controls on basis of design or analysis | a) **2 points: study controls for age and sex (or two other variables),** either by matching during patient and control recruitment or by adjusting for confounders in the analysis<br>b) **1 point: only one confounder** was controlled for<br>c) **0 points**: no significant difference between groups, but the confounders were not included in statistical analyses (e.g., age, and sex were not used as covariates in analysis and authors simply state there was no significant difference in those variables between groups) |
| **Exposure** | 1) Ascertainment of exposure | a) **1 point**: DTI data acquired; data processed and analyzed in blinded fashion<br>**Note: ALL studies received 1 point for this criterion** |
| | 2) Same method of ascertainment (i.e. DTI imaging) for cases and controls | a) **1 point**: **ALL studies received 1 point for this criterion** |
| | 3) Non-response rate | a) **1 point: no patient DTI data was excluded due to motion artifact or other reasons**, or an equal proportion of data was excluded for both patients and controls<br>b) **0 points**: inverse of the above statement |

**Table S4.** Assessment of study quality using the Newcastle-Ottawa Scale.

| Study | Quality Indices | | | | | | | | Total |
|---|---|---|---|---|---|---|---|---|---|
| | A | B | C | D | E | F | G | H | |
| Benedetti 2011 [1] | 1 | 0 | 0 | 1 | 2 | 1 | 1 | 1 | 7 |
| Besenek 2019 [2] | 1 | 0 | 0 | 1 | 2 | 1 | 1 | 1 | 7 |
| Cabeen 2018 [3] | 1 | 1 | 0 | 1 | 2 | 1 | 1 | 1 | 8 |
| Deng 2018 [4] | 1 | 0 | 0 | 1 | 2 | 1 | 1 | 0 | 6 |
| Dev 2017 [5] | 1 | 0 | 0 | 0 | 2 | 1 | 1 | 1 | 6 |
| Emsell 2014 [6] | 1 | 0 | 1 | 0 | 2 | 1 | 1 | 1 | 7 |
| Favre 2016 [7] | 1 | 0 | 1 | 1 | 2 | 1 | 1 | 1 | 8 |
| Foley 2018 [8] | 0 | 0 | 1 | 1 | 2 | 1 | 1 | 1 | 7 |
| Hermens 2018 [9] | 1 | 1 | 1 | 1 | 2 | 1 | 1 | 1 | 9 |
| Houenou 2007 [10] | 0 | 0 | 1 | 1 | 2 | 1 | 1 | 1 | 7 |
| Ji 2017 [11] | 1 | 1 | 1 | 1 | 2 | 1 | 1 | 1 | 9 |
| Knochel 2014 [12] | 1 | 0 | 0 | 1 | 2 | 1 | 1 | 1 | 7 |
| Koshiyama 2020 [13] | 1 | 0 | 1 | 1 | 2 | 1 | 1 | 0 | 7 |
| Lin 2011 [14] | 1 | 1 | 1 | 1 | 2 | 1 | 1 | 1 | 9 |
| Linke 2013 [15] | 0 | 1 | 1 | 1 | 2 | 1 | 1 | 1 | 8 |
| Linke 2020 [16] | 1 | 1 | 0 | 0 | 2 | 1 | 1 | 1 | 7 |
| Mahapatra 2017 [17] | 0 | 0 | 0 | 1 | 2 | 1 | 1 | 1 | 6 |
| Mamah 2019 [18] | 1 | 0 | 0 | 1 | 2 | 1 | 1 | 1 | 7 |
| McIntosh 2008 [19] | 1 | 1 | 1 | 0 | 2 | 1 | 1 | 1 | 8 |
| McKenna 2015 [20] | 1 | 0 | 1 | 1 | 2 | 1 | 1 | 1 | 8 |
| Moser 2018 [21] | 1 | 1 | 1 | 1 | 2 | 1 | 1 | 1 | 9 |
| Saricicek 2016 [22] | 1 | 0 | 0 | 1 | 2 | 1 | 1 | 1 | 7 |
| Sarrazin 2014 [23] | 1 | 1 | 1 | 1 | 2 | 1 | 1 | 1 | 9 |
| Scholz 2016 [24] | 1 | 0 | 1 | 1 | 2 | 1 | 1 | 1 | 8 |
| Shakeel 2020 [25] | 1 | 0 | 1 | 0 | 2 | 1 | 1 | 1 | 7 |
| Souza-Queiroz 2016 [26] | 1 | 1 | 1 | 1 | 2 | 1 | 1 | 0 | 8 |
| Sprooten 2013 [27] | 1 | 0 | 0 | 1 | 2 | 1 | 1 | 0 | 6 |
| Teixeira 2014 [28] | 1 | 0 | 1 | 1 | 2 | 1 | 1 | 1 | 8 |
| Tonnesen 2018 [29] | 1 | 1 | 1 | 0 | 2 | 1 | 1 | 0 | 7 |
| Tonnesen 2020 [30] | 0 | 1 | 0 | 0 | 2 | 1 | 1 | 0 | 5 |
| Versace 2014 [31] | 1 | 0 | 1 | 1 | 2 | 1 | 1 | 1 | 8 |
| Versace 2018 [32] | 1 | 0 | 1 | 1 | 2 | 1 | 1 | 0 | 7 |
| Yip 2013 [33] | 0 | 0 | 1 | 1 | 2 | 1 | 1 | 1 | 7 |

**Abbreviations: A,** Adequate definition of case; **B,** Representativeness of cases; **C,** Selection of control group; **D,** Definition of control group; **E,** Control for important confounding factors (e.g., age); **F,** Exposure assessment; **G,** Same method of ascertainment for cases and controls; **H,** Comparable number of cases and controls excluded due to motion

**Table S5.** Uncorrected p-values for null findings in inter-study heterogeneity.

| | Left UF | | Right UF | |
|---|---|---|---|---|
| | **β** | **p** | **β** | **p** |
| **Age** | 0.00 | 0.757 | -0.01 | 0.115 |
| **Sex** | 0.00 | 0.794 | 0.00 | 0.380 |
| **Lithium Use** | 0.00 | 0.987 | 0.00 | 0.682 |
| **Antipsychotic Use** | 0.00 | 0.528 | 0.00 | 0.922 |
| **Comorbid Anxiety** | 0.00 | 0.479 | 0.00 | 0.918 |
| **DTI Processing Pipeline** | 0.15 | 0.363 | 0.16 | 0.204 |

**Abbreviations: UF**, uncinate fasciculus; **DTI**, diffusion tensor imaging

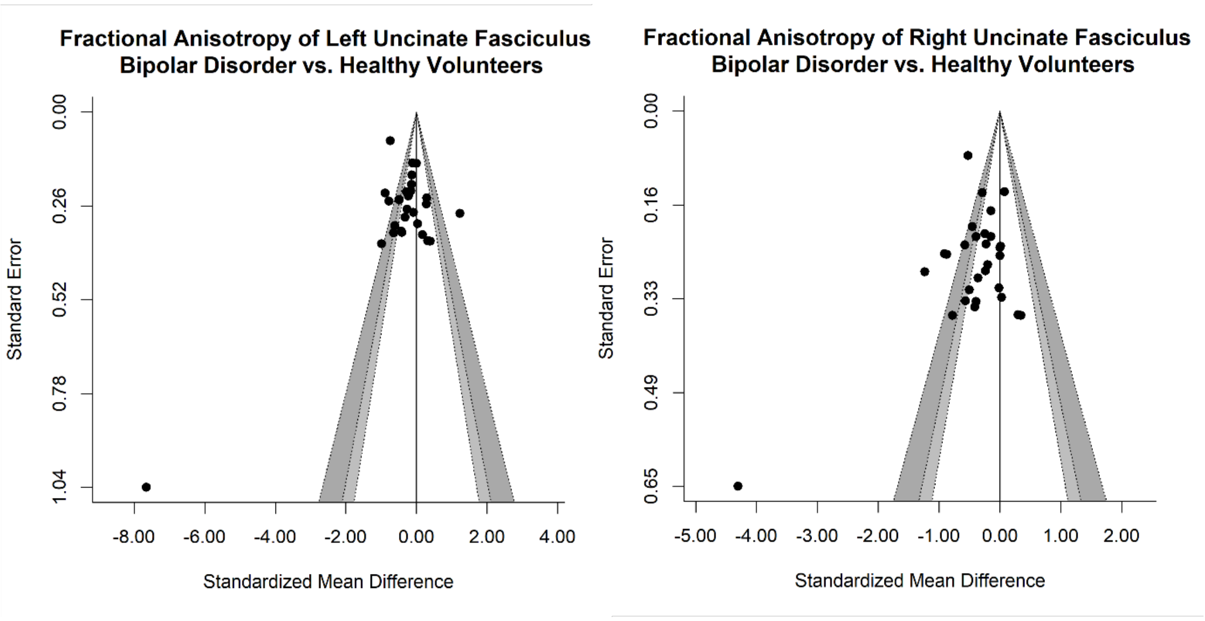


**Figure S1.** Funnel plot of fractional anisotropy in the left and right uncinate fasciculus of individuals with bipolar disorder versus healthy volunteers including the one outlier[17].

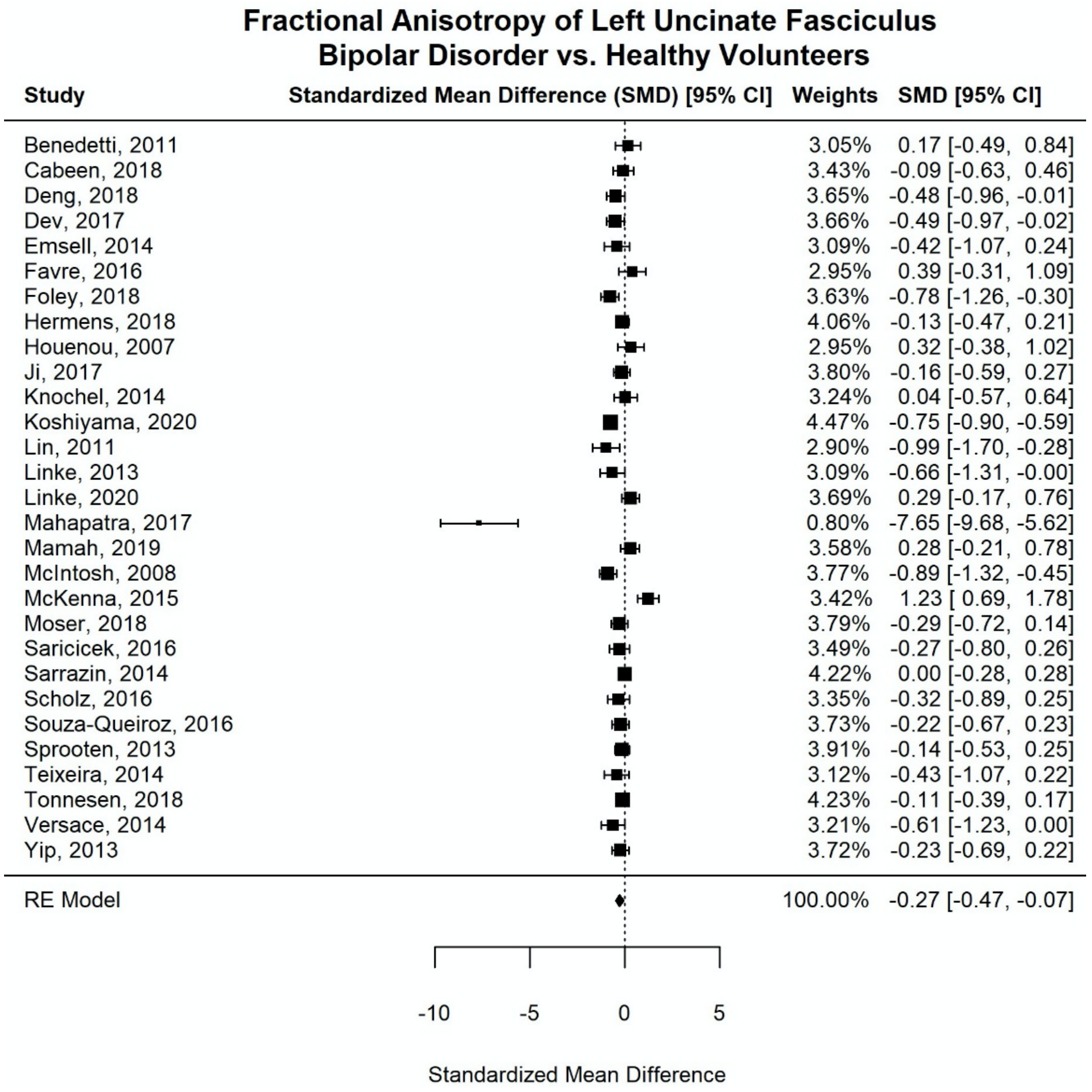


**Figure S2.** Forest plot of fractional anisotropy in the left uncinate fasciculus of individuals with bipolar disorder versus healthy volunteers including the one outlier[17].

**Abbreviations: CI**, confidence interval; **RE model**, random-effect model.

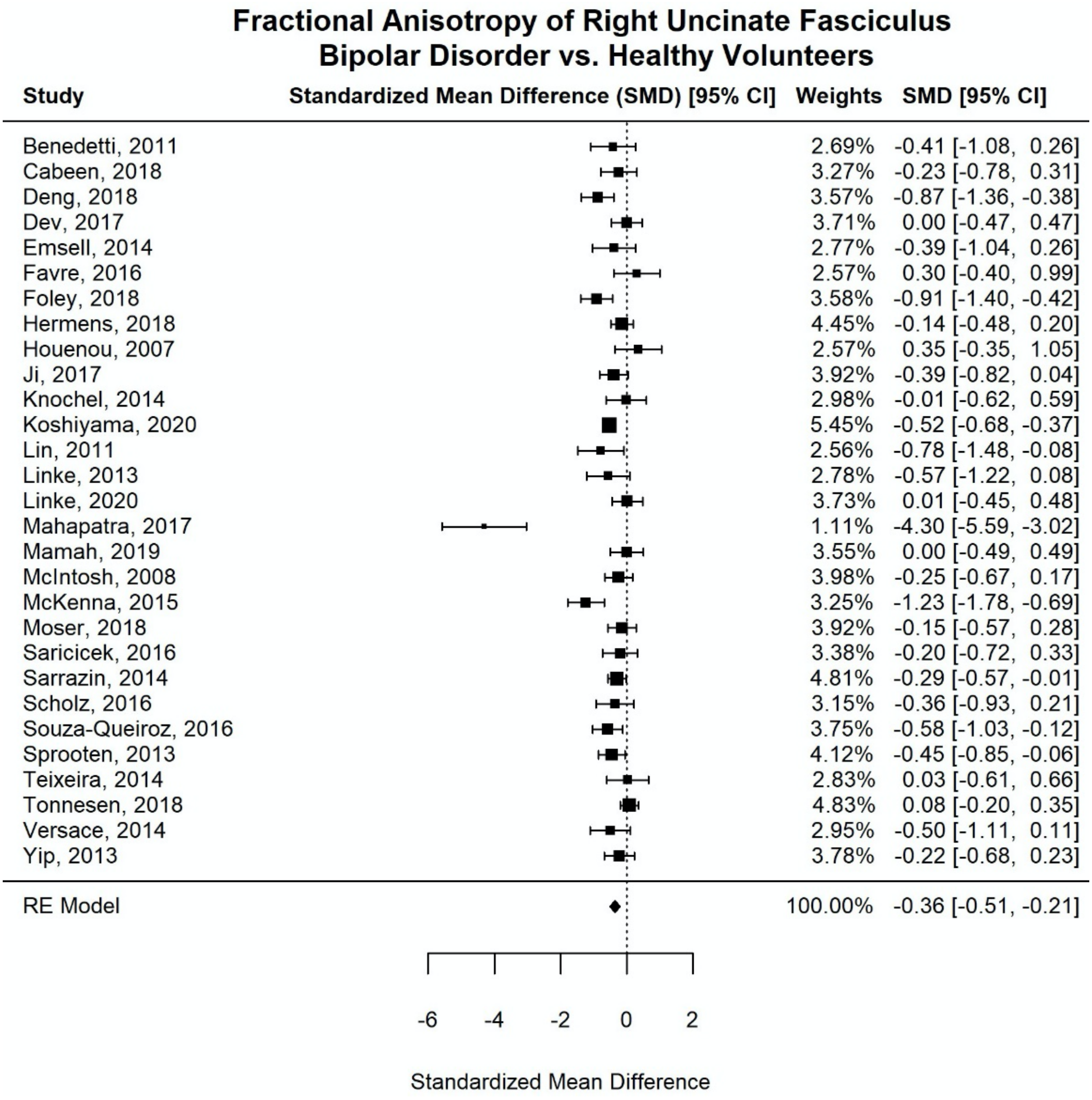


**Figure S3.** Forest plot of fractional anisotropy in the right uncinate fasciculus of individuals with bipolar disorder versus healthy volunteers including the one outlier[17].

**Abbreviations: CI**, confidence interval; **RE model**, random-effect model.

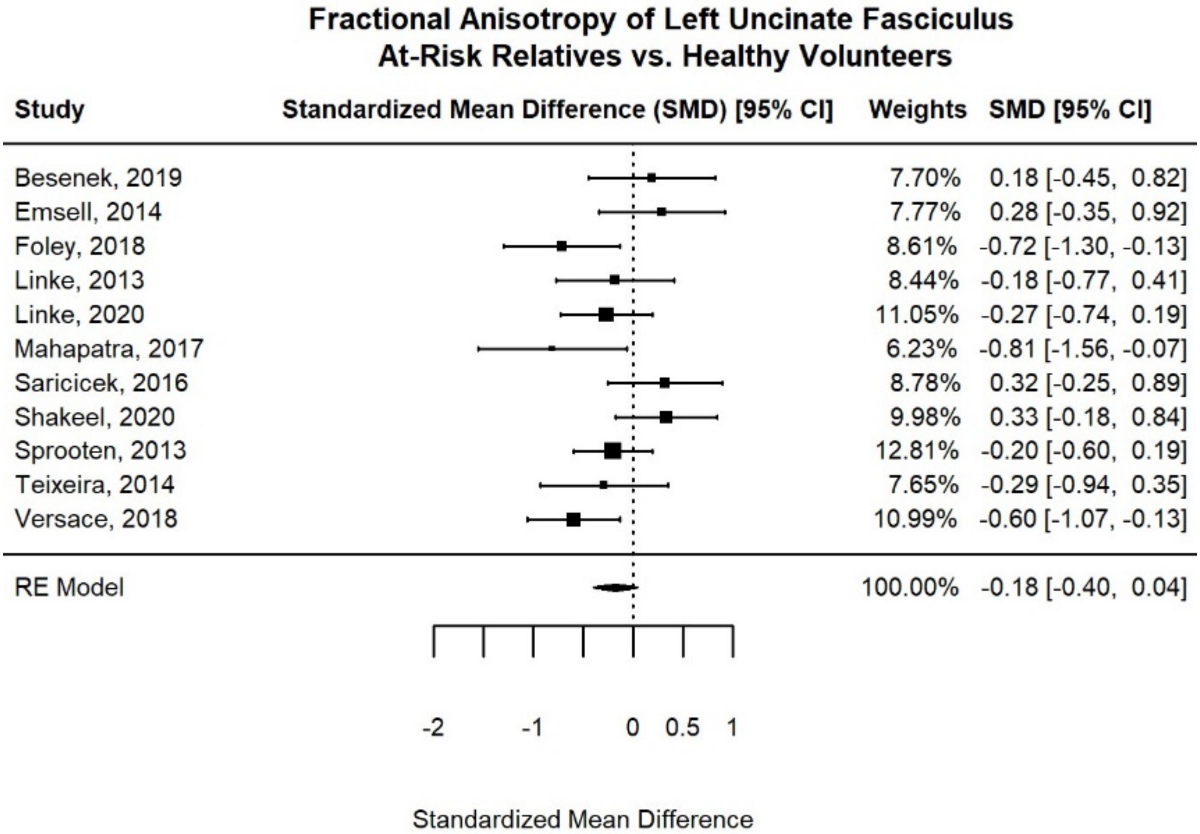


**Figure S4.** Forest plot of fractional anisotropy in the left uncinate fasciculus of at-risk relatives versus healthy volunteers.

**Abbreviations: CI**, confidence interval; **RE model**, random-effect model.

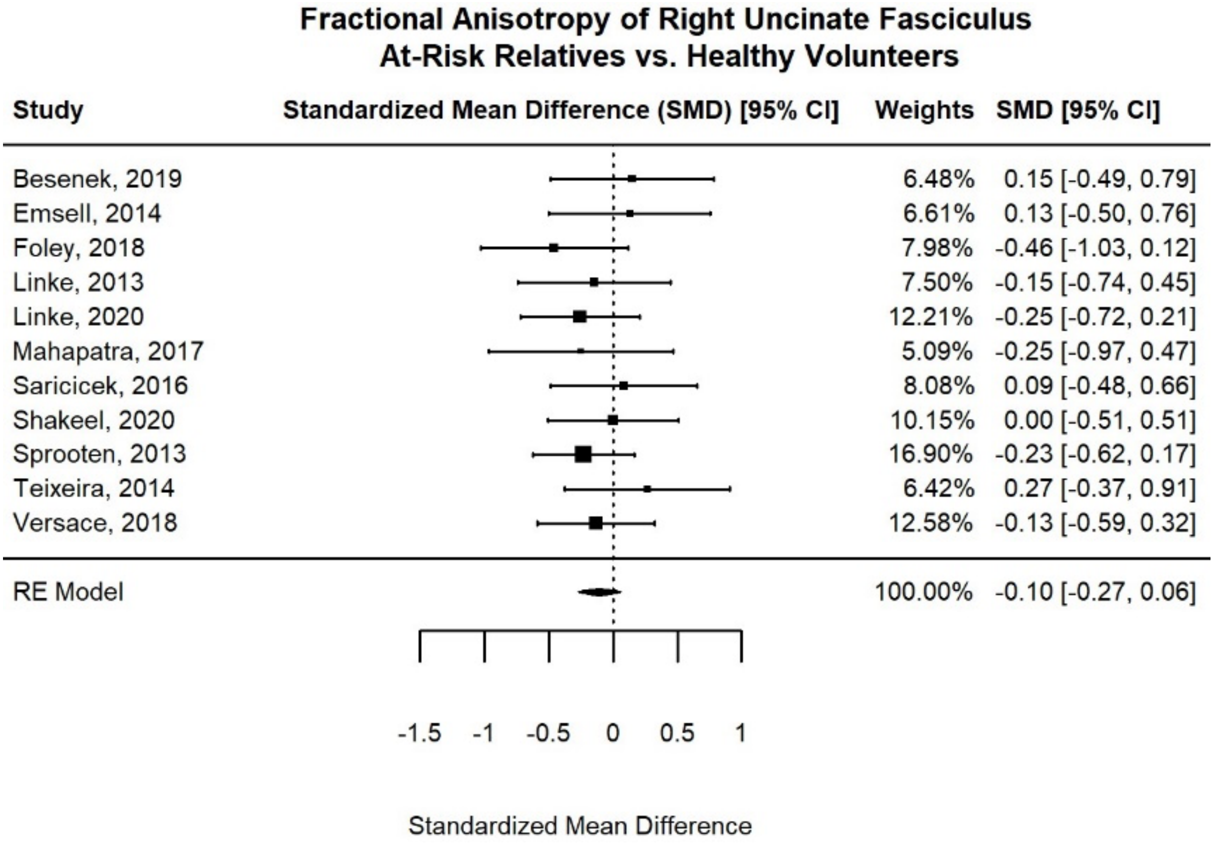


**Figure S5.** Forest plot of fractional anisotropy in the right uncinate fasciculus of at-risk relatives versus healthy volunteers.

**Abbreviations: CI**, confidence interval; **RE model**, random-effect model.

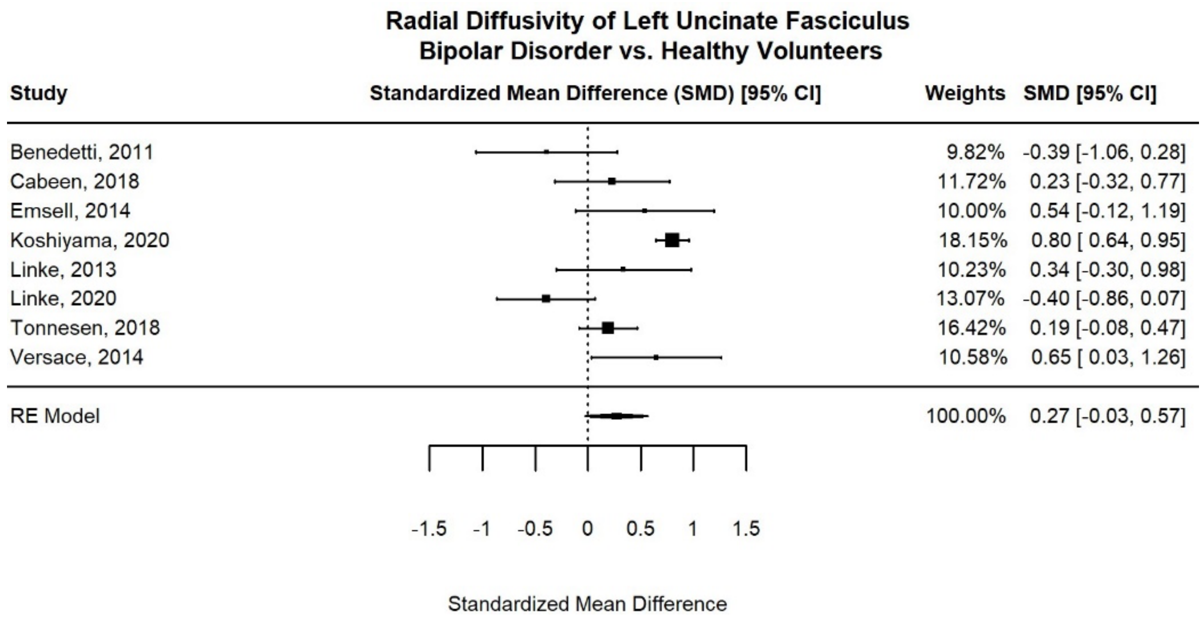


**Figure S6.** Forest plot of radial diffusivity in the left uncinate fasciculus of individuals with bipolar disorder versus healthy volunteers.

**Abbreviations: CI**, confidence interval; **RE model**, random-effect model.

relatives and healthy controls. *Psychiatry and Clinical Neurosciences*, *71*(10), 706–715. https://doi.org/10.1111/pcn.12530

26. Souza-Queiroz, J., Boisgontier, J., Etain, B., Poupon, C., Duclap, D., d'Albis, M.-A., Daban, C., Hamdani, N., Le Corvoisier, P., Delavest, M., Bellivier, F., Guevara, P., Leboyer, M., Henry, C., & Houenou, J. (2016). Childhood trauma and the limbic network: A multimodal MRI study in patients with bipolar disorder and controls. *Journal of Affective Disorders*, *200*, 159–164. https://doi.org/10.1016/j.jad.2016.04.038
27. Sprooten, E., Brumbaugh, M. S., Knowles, E. E. M., McKay, D. R., Lewis, J., Barrett, J., Landau, S., Cyr, L., Kochunov, P., Winkler, A. M., Pearlson, G. D., & Glahn, D. C. (2013). Reduced White Matter Integrity in Sibling Pairs Discordant for Bipolar Disorder. *American Journal of Psychiatry*, *170*(11), 1317–1325. https://doi.org/10.1176/appi.ajp.2013.12111462
28. Teixeira, A. M. A., Kleinman, A., Zanetti, M., Jackowski, M., Duran, F., Pereira, F., Lafer, B., Busatto, G. F., & Caetano, S. C. (2014). Preserved white matter in unmedicated pediatric bipolar disorder. *Neuroscience Letters*, *579*, 41–45. https://doi.org/10.1016/j.neulet.2014.06.061
29. Tønnesen, S., Kaufmann, T., Doan, N. T., Alnæs, D., Córdova-Palomera, A., Meer, D. van der, Rokicki, J., Moberget, T., Gurholt, T. P., Haukvik, U. K., Ueland, T., Lagerberg, T. V., Agartz, I., Andreassen, O. A., & Westlye, L. T. (2018). White matter aberrations and age-related trajectories in patients with schizophrenia and bipolar disorder revealed by diffusion tensor imaging. *Scientific Reports*, *8*(1), 14129. https://doi.org/10.1038/s41598-018-32355-9
30. Tønnesen, S., Kaufmann, T., Lange, A.-M. G. de, Richard, G., Doan, N. T., Alnæs, D., Meer, D. van der, Rokicki, J., Moberget, T., Maximov, I. I., Agartz, I., Aminoff, S. R., Beck, D., Barch, D. M., Beresniewicz, J., Cervenka, S., Fatouros-Bergman, H., Craven, A. R., Flyckt, L., … Westlye, L. T. (2020). Brain Age Prediction Reveals Aberrant Brain White Matter in Schizophrenia and Bipolar Disorder: A Multisample Diffusion Tensor Imaging Study. *Biological Psychiatry: Cognitive Neuroscience and Neuroimaging*, *5*(12), 1095–1103. https://doi.org/10.1016/j.bpsc.2020.06.014
31. Versace, A., Andreazza, A. C., Young, L. T., Fournier, J. C., Almeida, J. R. C., Stiffler, R. S., Lockovich, J. C., Aslam, H. A., Pollock, M. H., Park, H., Nimgaonkar, V. L., Kupfer, D. J., & Phillips, M. L. (2014). Elevated serum measures of lipid peroxidation and abnormal prefrontal white matter in euthymic bipolar adults: Toward peripheral biomarkers of bipolar disorder. *Molecular Psychiatry*, *19*(2), 200–208. https://doi.org/10.1038/mp.2012.188
32. Versace, A., Ladouceur, C. D., Graur, S., Acuff, H. E., Bonar, L. K., Monk, K., McCaffrey, A., Yendiki, A., Leemans, A., Travis, M. J., Diwadkar, V. A., Holland, S. K., Sunshine, J. L., Kowatch, R. A., Horwitz, S. M., Frazier, T. W., Arnold, L. E., Fristad, M. A., Youngstrom, E. A., … Phillips, M. L. (2018). Diffusion imaging markers of bipolar versus general

psychopathology risk in youth at-risk. *Neuropsychopharmacology*, *43*(11), 2212–2220. https://doi.org/10.1038/s41386-018-0083-z

33. Yip, S. W., Chandler, R. A., Rogers, R. D., Mackay, C. E., & Goodwin, G. M. (2013). White matter alterations in antipsychotic- and mood stabilizer-naïve individuals with bipolar II/NOS disorder. *NeuroImage: Clinical*, *3*, 271–278. https://doi.org/10.1016/j.nicl.2013.08.005
